# Comparing an AI test to a 21-gene assay for premenopausal node-positive HR+/HER2-breast cancer

**DOI:** 10.64898/2026.02.06.26345771

**Authors:** Jailan Elayoubi, Cerise Tang, Kathryn J. Ruddy, Khalil Choucair, Kevin Kalinsky, Katarzyna Pogoda, Francisco J. Esteva, Jad M Abdelsattar, Virginia F Borges, Ken Zeng, Joseph Cappadona, Bartosz Machura, Dhruva Biswas, Krzysztof J. Geras, Jan Witowski

## Abstract

Recurrence scores based on a 21-gene assay are clinically useful for predicting prognosis and chemotherapy benefit in postmenopausal node-positive breast cancer patients, but its performance in premenopausal patients is inconsistent. Here, we evaluated Ataraxis Breast RISK (ATX), an AI test that predicts recurrence risk, and compared it with the genomic assay. ATX identified high risk patients misclassified as low risk by the genomic assay and therefore may refine selection of patients for adjuvant chemotherapy.

## Main

Approximately one third of women diagnosed with hormone receptor (HR)-positive, human epidermal growth factor receptor 2 (HER2)-negative breast cancer present with lymph-node-positive disease, which is associated with higher rates of recurrence.^1,2^ The RxPONDER trial^3^ was a landmark study demonstrating that the 21-gene Oncotype DX (ODX) assay^4,5^ can help guide adjuvant chemotherapy use in postmenopausal women with 1-3 positive lymph nodes and an ODX recurrence score ≤25. However, in premenopausal women, interpretation of ODX recurrence scores is complicated by the ovarian-suppressive effects of chemotherapy, making it difficult to distinguish which patients truly benefit from cytotoxic therapy versus chemotherapy induced amenorrhea. Therefore, current NCCN guidelines recommend ODX only as a consideration in this patient subgroup, leaving risk-adapted decision-making unresolved and up to the discretion of their physician.

In this study, we used Ataraxis Breast RISK (ATX),^6^ a multimodal artificial intelligence (AI) test that integrates clinical data with morphological features extracted from H&E-stained primary tumor slides, to validate its prognostic accuracy in premenopausal, node-positive women. We assembled a cohort of 150 premenopausal node-positive HR+/HER2-patients across 4 institutions with available H&E-stained slides and complete clinical information (**Table 1**). The median follow-up within this cohort was 4.75 years (reverse Kaplan-Meier estimate)^7^ and across the full follow-up period, 20 recurrence events were observed (event rate = 13.3%). ATX generated a predicted probability of breast cancer recurrence within 5 years (**Extended Data Figure 1**). A threshold of 10% risk of recurrence was used to stratify patients into ATX high (score greater or equal to 10%) or ATX low (less than 10%) risk groups. Within the 150-patient cohort, 111 patients were classified as ATX high and 39 patients were classified as ATX low.

**Table 1:** Characteristics of 150 premenopausal women in our evaluation cohort. Continuous variables are presented as mean (minimum-maximum), and categorical variables are presented as n (% of patients).

| Characteristic | Category | Summary |
| --- | --- | --- |
| Grade | 1.0 | 12 (11.9%) |
|  | 2.0 | 62 (61.4%) |
|  | 3.0 | 27 (26.7%) |
| T stage | T1b | 3 (2.0%) |
|  | T1c | 52 (34.7%) |
|  | T2 | 73 (48.7%) |
|  | T3 | 21 (14.0%) |
|  | T4 | 1 (0.7%) |
| N Stage | N1 | 50 (34.0%) |
|  | N1a | 44 (29.9%) |
|  | N1mi | 20 (13.6%) |
|  | N2 | 7 (4.8%) |
|  | N2a | 16 (10.9%) |
|  | N3 | 6 (4.1%) |
|  | N3a | 4 (2.7%) |
| Age at Diagnosis |  | 43.3 (27-50) |
| Adjuvant Endocrine Therapy | 0.0 | 9 (11.5%) |
|  | 1.0 | 69 (88.5%) |
| Adjuvant Chemotherapy | 0.0 | 30 (39.0%) |
|  | 1.0 | 47 (61.0%) |
| ATX Risk Score |  | 0.2 (0-1) |
| ATX Risk Category | Low | 39 (26.0%) |
|  | High | 111 (74.0%) |

We examined the statistical association between ATX scores and recurrence-free interval (RFI), which served as the primary endpoint in this study. Among 150 premenopausal node-positive patients, ATX risk groups showed a significant difference in KM-estimated probability of RFI (p_log-rank_ = 0.04) (**Figure 1A**). Specifically, ATX high patients had a lower 5-year probability of remaining recurrence-free (84%, 95% CI = 73%-90%) than ATX low patients (91%, 95% CI = 68%-98%). Notably, for the first four years of follow-up, 100% of patients identified as ATX low risk did not experience an RFI-contributing event. We further considered the possibility that other clinical factors may confound an association between ATX scores and RFI. To directly examine this probability, we used a multivariate Cox model stratified by dataset and adjusted for tumor stage, grade, and age at diagnosis. ATX score (HR per 0.1 increase in ATX score) remained independently associated with RFI (HR = 6.10, 95% CI = 1.45-25.31, p = 0.01), whereas no other clinical variable reached significance, though effect size estimates were imprecise (**Figure 1B**). These findings indicate that ATX provides prognostic information beyond established clinical factors, supporting its potential as an independent, AI-enhanced predictor of recurrence risk in HR+/HER2-premenopausal node-positive breast cancer patients.

**Figure 1:**
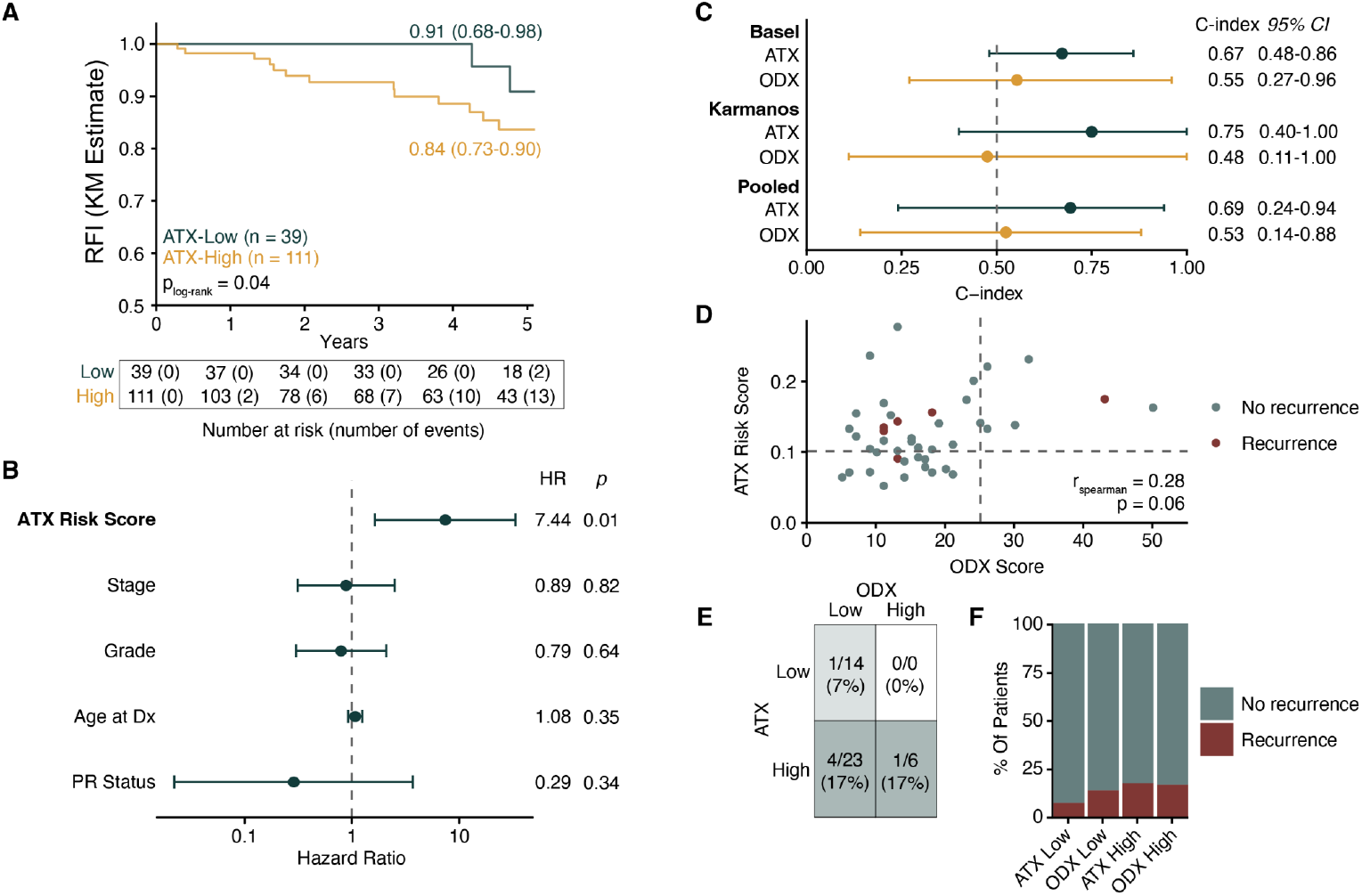
Prognostic performance of ATX versus ODX in premenopausal node-positive HR+/HER2– breast cancer. a) Kaplan-Meier estimates of 5-year RFI in 150 patients stratified by ATX risk score (cut-off = 0.1). p_log-rank_ represents the log-rank test p-value. RFI at 5-years and corresponding 95% confidence intervals are shown. b) Multivariate Cox regression demonstrating that ATX risk score (per 0.1 increase) is associated with RFIindependent of other clinical factors in 150 patients. Wide confidence intervals suggest directional robustness but low precision in estimates. c) Prognostic discrimination of ATX versus Oncotype DX (ODX) scores in the paired subset (n=43), quantified by Harrell’s concordance index (C-index). Cohort-specific estimates were pooled using a random effects model, with higher concordance for ATX. d) Scatterplot comparing ATX and ODX scores. Several patients classified as low risk by ODX were designated as high risk by ATX. e) Heatmap of 5-year recurrence rates stratified by ATX and ODX risk groups. Among ODX low patients (n=37), 23 were ATX high, with 4 (17%) recurring within 5 years. f) Stacked barplot showing the proportion of patients with and without 5-year recurrence across risk categories. Recurrence enrichment was greater for ATX high vs low (17% vs 7%), whereas recurrence proportions were comparable between ODX groups (17% vs 14%).

To compare the performance of ATX to Oncotype DX, we narrowed our analyses to patients who had ODX recurrence scores available (n = 43). We computed Harrell’s C-indices^8–10^ within each cohort and subgroup metrics were pooled using a random effects model. Across cohorts, both ATX and ODX demonstrated discrimination for recurrence-free interval, where ATX showed higher concordance (C-index = 0.69, 95% CI = 0.24-0.94) than ODX (C-index = 0.53, 95% CI = 0.14-0.88 (**Figure 1C**). However, the small sample size of this cohort led to imprecise estimates with wide confidence intervals. Comparing ATX scores and ODX scores, we found they were not significantly correlated (r = 0.28, p = 0.06). 14 patients were concordantly classified as low risk (ODX ≤25) and 6 were high risk across both scores (**Figure 1D**). However, 23 patients were classified as low risk by ODX but high risk by ATX, suggesting classification by ODX only may miss patients who could benefit from adjuvant chemotherapy. Among the 23 discordant patients classified as ODX low but ATX high, 13 did not receive adjuvant chemotherapy. Within this group, 3 of 13 (23%) experienced recurrence within 5 years, compared with 1 of 10 (10%) among those who received chemotherapy. Although underpowered for statistical comparison, this trend suggests ATX may identify patients who could benefit from adjuvant chemotherapy despite low genomic risk classification. No patients were classified as ATX low + ODX high.

At five years, recurrence events were observed in 7% of patients classified as ATX low + ODX low compared to 17% of ATX high + ODX high patients. Notably, the discordant ATX high + ODX low group also demonstrated a recurrence rate of 17%, with 4 out of 23 patients experiencing an event (**Figure 1E**). Overall, 5 of 29 (17%) of ATX high patients recurred, compared to 1 out of 14 (7%) of ATX low patients, indicating strong discriminatory ability (**Figure 1F**). In contrast, ODX high and ODX low patients had comparable 5-year recurrence rates (17% vs 14%). Notably, most recurrence events (4 out of 6) occurred in the discordant ODX low + ATX high group, underscoring ATX’s potential ability to identify aggressive disease not recognized by genomic scoring, highlighting a population that may benefit from alternative treatment strategies despite low genomic risk scores.

Within patients classified as ATX low, outcomes were overwhelmingly favorable. Only 1 of 14 (7%) experienced a recurrence event within 5 years. Notably, none of the 4 ATX low patients who received adjuvant chemotherapy recurred, and among the 10 who did not receive adjuvant chemotherapy, 9 (90%) remained disease-free at 5 years. These findings parallel results from the SWOG 8814 and RxPONDER trials, where Oncotype DX identified postmenopausal low risk patients who could safely avoid chemotherapy. ATX appears to be capable of extending this finding to premenopausal node-positive patients by identifying a group with excellent outcomes despite limited therapy, a level of discrimination that was not consistently achieved by previous genomic assays.

Previous work has demonstrated that while genomic assays effectively predict the benefit of adjuvant chemotherapy in postmenopausal HR+/HER2-node positive breast cancers, their utility in premenopausal patients is limited. In trials such as RxPONDER, the apparent benefit of chemotherapy in premenopausal women may reflect chemotherapy induced amenorrhea rather than direct cytotoxic effect, leaving unclear which patients require treatment escalation. Our results confirm these observations, showing that premenopausal node-positive patients experience similar rates of 5-year recurrence regardless of Oncotype DX risk category. These findings underscore the need for alternative approaches to risk stratification that leverage routinely available data.

Our data suggest that Ataraxis Breast RISK, a multimodal AI test integrating histology images with clinical data, is able to stratify premenopausal node-positive HR+/HER2-breast cancer patients by recurrence risk and thus inform both treatment escalation and de-escalation. ATX leverages a cutting-edge vision-transformer based foundation model trained on over 400 million pathology image slides using self-supervised learning.^11^ Importantly, ATX derives prognostic information directly from routinely collected histopathology slides, without the use of costly molecular profiling. This positions it as a scalable tool whereas access to multigene assays may remain limited by cost or technical constraints. Beyond scalability, our findings suggest that histology-informed deep learning tests, such as ATX, may capture features of tumor biology not represented in transcriptomic panels. The ability of vision transformer-based architectures to interpret spatial context and microenvironmental cues may explain ATX’s stronger performance.

While the results presented are promising, as a multisite observational analysis of premenopausal women with HR+/HER2-early breast cancer, our study has several limitations. First, the sample size is modest and we were underpowered to detect statistical differences between ATX and ODX risk groups. Second, effect-size uncertainty is substantial, as illustrated with wide CIs, due to event scarcity and potential upper-tail leverage and therefore estimates should be interpreted as directionally robust but imprecise. Third, while two datasets had both ATX and ODX scores, allowing us to demonstrate generalizability of the Ataraxis test, both cohorts were retrospectively assembled. Fourth, cause-specific mortality was unavailable for all patients and thus RFI in this study is an approximation and not a strict implementation of the STEEP criteria. Lastly, limited information on chemotherapy, type of hormonal therapy, and ovarian function suppression treatment restricts our ability to draw conclusions on true treatment effects.

In the future, prospective validation studies of ATX are planned to confirm clinical utility. Moreover, post-deployment surveillance will be required to establish its clinical utility and integration in routine care. Finally, as with all AI tests, model interpretability linking morphological patterns with biological processes is important. Future explainability studies will clarify the biological mechanisms underpinning the risk signal and could further support clinical adoption.

Our findings highlight the potential for large-scale, image-based artificial intelligence tests to be used in breast cancer. Ataraxis Breast RISK demonstrates promising independent prognostic performance in premenopausal node-positive breast cancer patients across diverse datasets. By enabling risk assessment directly from routinely-collected pathology slides, ATX offers a scalable and accessible alternative to molecular assays and may help refine adjuvant chemotherapy treatment decisions for a patient population in whom genomic tests are not prognostic. Importantly, ATX refines risk stratification at both extremes, identifying premenopausal patients with node-positive HR+/HER2-early breast cancer harboring residual recurrence risk missed by genomic testing, as well as a smaller group with excellent outcomes who may be eligible for therapy de-escalation.

## Methods

### Patient cohort

Patient cases with H&E-stained histology images and complete clinical information were collected from four institutions. In total, 150 premenopausal HR+/HER2-node-positive cases were extracted from TCGA (n=59), Providence Health (n=48), Basel (n=32), and Karmanos (n=11). Of these 150 patients, 43 had Oncotype DX scores available (Basel n=32, Karmanos n=11).

Among the 150 patients, adjuvant chemotherapy information was available for 77 individuals, of whom 47 received chemotherapy and 30 did not. Adjuvant endocrine therapy information was available for 78 patients; 69 received endocrine therapy, whereas 9 did not. Within the subset of patients with Oncotype DX scores, adjuvant chemotherapy information was available for 42 patients, including 18 who received chemotherapy and 24 who did not. Adjuvant endocrine therapy information was available for all patients in this subset, with 41 receiving endocrine therapy.

### Development and training of Ataraxis Breast RISK

The Ataraxis Breast RISK (ATX) test processes digitized H&E slide images and integrates it with clinical data including patient’s age, tumor and nodal stage, ER status, PR status, HER2 status, and histological subtype.^11^ Morphological features from the slide are extracted using Kestrel, a vision transformer-based model trained on 400 million pathology images.^22^ Morphological features are then extracted and integrated with clinical variables to predict the probability of recurrence at 5 years. ATX was trained as a time-to-event model on over 4,500 samples distinct from the samples used in this validation study. A higher ATX score suggests a higher probability of recurrence. ATX is a class C1 prognostic biomarker based on 2025 European Society of Medical Oncology (ESMO) biomarker classification criteria.^12^

### Survival analysis

The primary endpoint used was recurrence-free interval (RFI), defined as duration from diagnosis to first recurrence, excluding death as cause-specific mortality was unavailable for patients in this study. Risk stratification and discriminatory ability were measured by Harrell’s concordance index.^8–10^ 95% confidence intervals were obtained using 1000 bootstrap iterations. Hazard ratio was used to measure relative change in risk of recurrence with every 0.1 unit increase in score. Hazard ratios were calculated using multivariate Cox models.

Kaplan-Meier curves were estimated using the KaplanMeierFitter class from lifelines,^13^ confidence intervals were calculated using Greenwood’s formula, and p-values were calculated using log-rank tests. CoxPHFitter class from lifelines was used to fit the multivariate Cox proportional hazards model and clinical features included were tumor stage, grade, and age at diagnosis. Tumor stage and grade were treated as ordinal variables while age was modeled as a continuous variable. All 150 patients had complete tumor stage and age data, while 49 patients had no grade data. Median follow up time was calculated using the reverse Kaplan-Meier method.

### Meta-analysis

Concordance indices were pooled using random effects models and were performed using the R meta package (v8.2-1)^14^ and assessed via rpy2.

### Data visualization

All figures were made using matplotlib^15^ and ggplot2.^16^

### Statistics and reproducibility

Sample sizes were not predetermined using statistical methods. Statistical analyses were performed using Python 3.10 and R4.0+ via rpy2. Data management and manipulation in Python were conducted using pandas (v1.5)^17^ and numpy.^18^

## Extended Data

**Extended Data Figure 1:**
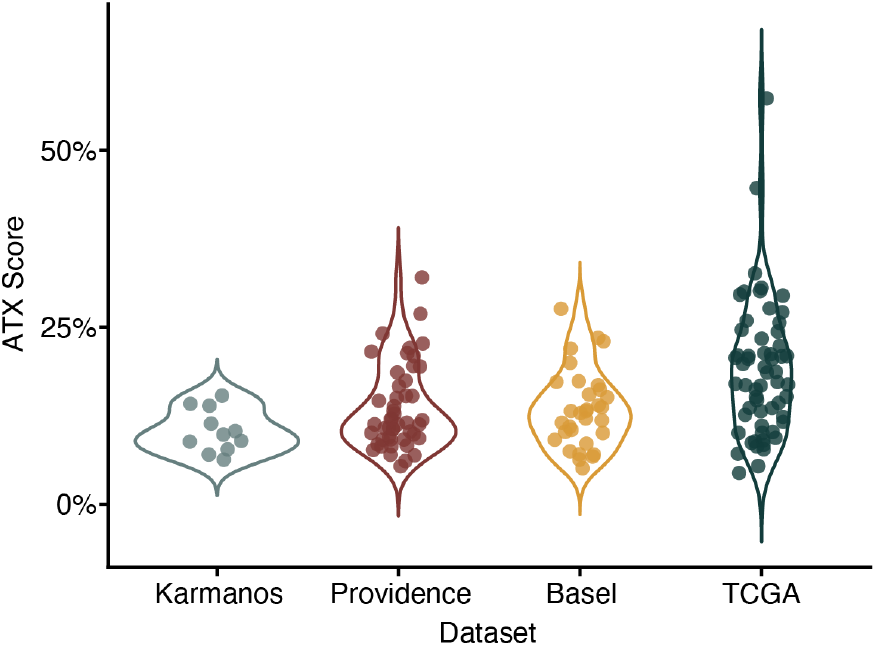
Distribution of ATX scores across datasets. ATX scores across 4 breast cancer datasets. Each dot corresponds to a patient. The y-axis represents the patient’s ATX score. Datasets are ordered by median ATX score, with the lowest score on the left and the highest score on the right.

## Data Availability

Data from The Cancer Genome Atlas (TCGA) are available via The Cancer Imaging Archive (TCIA) under DOI: 10.7937/K9/TCIA.2016.AB2NAZRP. The Providence data is available through Nightingale under DOI: 10.48815/N5159B. All other datasets analyzed in this study are private or proprietary and are not publicly available.

## Code Availability

Ataraxis Breast RISK can be accessed for non-commercial purposes. A jupyter notebook to replicate analyses is available upon request.

## Acknowledgements

This study makes use of data generated by the Molecular Taxonomy of Breast Cancer International Consortium. Funding for the project was provided by Cancer Research UK and the British Columbia Cancer Agency Branch. We also acknowledge the primary METABRIC publication.^38^ Tissues and samples were received from the Australian Breast Cancer Tissue Bank which is generously supported by the National Health and Medical Research Council of Australia, The Cancer Institute NSW and the National Breast Cancer Foundation. The tissues and samples are made available to researchers on a non-exclusive basis. The authors wish to acknowledge the roles of the Barts Cancer Now Tissue Bank in collecting and making available the samples and/or data, and the patients who have generously donated their tissues and shared their data to be used in the generation of this publication. Biosamples were obtained from the Wales Cancer Bank (DOI: http://doi.org/10.5334/ojb.46) which is funded by Health and Care Research Wales. Other investigators may have received specimens from the same subjects. We acknowledge the contributions (contributed materials) of the Northern Ireland Biobank in all academic outputs incorporating, or resulting from the use of, the material. Biological materials were provided by the Ontario Tumour Bank, which is supported by the Ontario Institute for Cancer Research through funding provided by the Government of Ontario. The views expressed in this publication are the views of the authors and do not necessarily reflect those of the Government of Ontario. The results published here are in part based upon data generated by the TCGA Research Network: http://cancergenome.nih.gov/.

## Author Contributions

JE, KJG, and JW conceived the study. JE, CT, JC, KZ, KJG, JW assisted with data collection and analytical methodology development. JE, CT, KJG, and JW designed and performed the experiments. JE, CT, KJG, and JW wrote the paper. JE, CT, KJR, KC, KK, KP, FJE, JMA, VB, KZ, JC, BM, DB, KJG, and JW critically reviewed the manuscript and provided final approval.

